# The effectiveness of non-pharmaceutical interventions in containing epidemics: a rapid review of the literature and quantitative assessment

**DOI:** 10.1101/2020.04.06.20054197

**Authors:** Jane Cheatley, Sabine Vuik, Marion Devaux, Stefano Scarpetta, Mark Pearson, Francesca Colombo, Michele Cecchini

## Abstract

The number of confirmed COVID-19 cases has rapidly increased since discovery of the disease in December 2019. In the absence of medical countermeasures to stop the spread of the disease (i.e. vaccines), countries have responded by implementing a suite of non-pharmaceutical interventions (NPIs) to contain and mitigate COVID-19. Individual NPIs range in intensity (e.g. from lockdown to public health campaigns on personal hygiene), as does their impact on reducing disease transmission. This study uses a rapid review approach and investigates evidence from previous epidemic outbreaks to provide a quantitative assessment of the effectiveness of key NPIs used by countries to combat the COVID-19 pandemic. Results from the study are designed to help countries enhance their policy response as well as inform transition strategies by identifying which policies should be relaxed and which should not.

## Introduction

In December 2019, Wuhan, located in the Hubei province of China, experienced an outbreak of pneumonia from a novel virus – severe acute respiratory syndrome coronavirus 2 (SARS-CoV-2) (Chen et al., 2020_[1]_). SARS-CoV-2 can lead to the coronavirus disease 2019 (i.e. COVID-19), which has symptoms ranging from a cough or fever, to more severe illnesses such as pneumonia and respiratory stress, which may result in death (World Health Organization, 2020_[2]_).

The number of confirmed COVID-19 cases has grown rapidly since December and spread to countries across the world. Reasons for disease’s rapid proliferation include: the large number of people experiencing no or mild symptoms (Xu et al., 2020_[5]_); the disease’s relatively long incubation period (Lauer et al., 2020_[6]_; Baum, 2020_[7]_; World Health Organization, 2018_[8]_); a high reproduction number (Biggerstaff et al., 2014_[9]_; World Health Organization, 2020_[10]_; Wang et al., 2020_[11]_); and the capacity for the virus to last on surfaces for up to three days (van Doremalen et al., 2020_[12]_).

In the absence of medical countermeasures to prevent the spread of the disease, policy-makers are reliant on non-pharmaceutical interventions (NPIs) to both contain and mitigate its impact. The former relates to strategies that minimise the risk of transmission (i.e. reducing the reproduction number to below one), while the latter aims to slow the disease’s progress and lessen its impact. Although containment and mitigation strategies have different objectives, their interventions largely overlap and are often implemented concurrently. Further, they both work to alleviate the burden on health care systems by ‘buying time’ for patients occupying hospital beds to recover, and by keeping the number of new patients to a manageable level.

This study provides a quantitative assessment of the potential effectiveness of key NPIs utilised by countries to combat the COVID-19 outbreak, namely social distancing, school closures, travel restrictions, contact tracing and quarantine, public information campaigns, and environmental and personal hygiene measures. Findings from the study are designed to assist policy-makers adjust, if necessary, their strategy to fighting the outbreak, by outlining the effectiveness of NPIs, in isolation or as part of a comprehensive policy package, on key outcome measures. Once the number of new cases falls and the impact of the disease lessens, the findings can also be used to inform a country’s transition strategy, for example, by identifying which interventions to relax and which to not.

## Methodology

A rapid review of the literature was undertaken to identify salient NPIs used to combat influenza outbreaks and their associated effectiveness. Academic, peer-reviewed articles were sourced from major databases including those focused on public health – e.g. MEDLINE, and Google Scholar. The research was carried out on 19 March 2020.

The search for articles concentrated on a combination of key terms including ‘influenza’, ‘pandemic’, ‘non-pharmaceutical measures’, ‘non-pharmaceutical interventions’, ‘social distancing’, ‘school closures’, ‘workplace closures’, ‘mass gatherings’, ‘hygiene’, ‘contact tracing’, ‘travel restrictions’, ‘quarantine’ and ‘public health campaigns’. Additional articles were sourced using a snowball approach.

To the extent possible, systematic reviews and meta-analyses pertaining to the effectiveness of NPIs were referenced in order to provide an up-to-date summary of the literature. To compare the relative effectiveness of policies, two common outcome measures frequently cited in the literature were identified (i.e. change in the overall attack rate and delay in the peak of disease). Data on these two dimensions was collected from the reviewed papers and compared using standard graphical approaches. Data on other dimensions was also collected and reported in the paper, however, a systematic comparison was not possible due to heterogeneity in the measures used.

Finally, policies being utilised by countries to combat COVID-19 were obtained from the grey literature, in particular official government websites and documents.

## Results

### Social distancing

Social distancing refers to policies that deliberately increase physical space between people (John Hopkins Medicine, 2020_[1]_). Such policies come in many forms including banning large gatherings; encouraging people to work from home; and closure of non-essential stores such as restaurants and cafes. These policies may be implemented across a community or target specific at-risk groups such as the elderly and those with pre-existing health conditions (Anderson et al., 2020_[2]_). The primary objective of social distancing is to prevent transmission thereby flattening the peak of the disease (Anderson et al., 2020_[2]_). In the short-term this will ease pressure on the health care system, while in the long-term it provides time for new treatment and vaccines to be developed (Anderson et al., 2020_[2]_).

Several studies analysing the impact of social distancing on disease outbreaks exist (Chen et al., 2020_[3]_). For example, a systematic review of workplace social distancing found the policy reduces the influenza attack rate (i.e. the proportion of individuals in a population who contract the disease) by 23% in the general population (Ahmed, Zviedrite and Uzicanin, 2018_[4]_). This is supported by an earlier study which found working-from-home was ‘moderately effective’ in reducing influenza transmission by 20-30% (Rashid et al., 2015_[5]_). Further, social distancing in Sydney, Australia, during the 1918-19 influenza pandemic is estimated to have reduced the attack rate by nearly 40% (i.e. from 60% to 37%) indicating 22% of the population were spared infection (Caley, Philp and McCracken, 2008_[6]_).

One approach to social distancing is the banning of mass events (e.g. music festivals or large spectator sporting events). While often seen as a logical element of containment strategies, the evidence suggest that this intervention is most effective when implemented together with other social distancing measures (Ishola and Phin, 2011_[7]_; Markel et al., 2007_[8]_). This is because contact-time at such events is relatively small compared to the time spent in schools, workplaces, or other community locations such restaurants (Ferguson et al., 2020_[9]_). As with other containment strategies, the earlier bans on mass gatherings are enforced, the greater their impact (Hatchett, Mecher and Lipsitch, 2007_[10]_).

Several challenges are associated with social distancing. Salient examples include reduced economic activity caused by closures and reduced social interaction (Rashid et al., 2015_[5]_; OECD, 2020_[11]_); neglect of vulnerable populations, such as the elderly (Boddy, Young and O’Leary, 2020_[12]_); and psychological damage such as acute distress disorder, anxiety and insomnia (Brooks et al., 2020_[13]_).

Table 1 lists certain policies implemented by OECD countries to enhance social distancing in response to the COVID-19 pandemic. Examples range in their intensity and thus their impact on day-to-day life for affected populations.

**Table 1.**
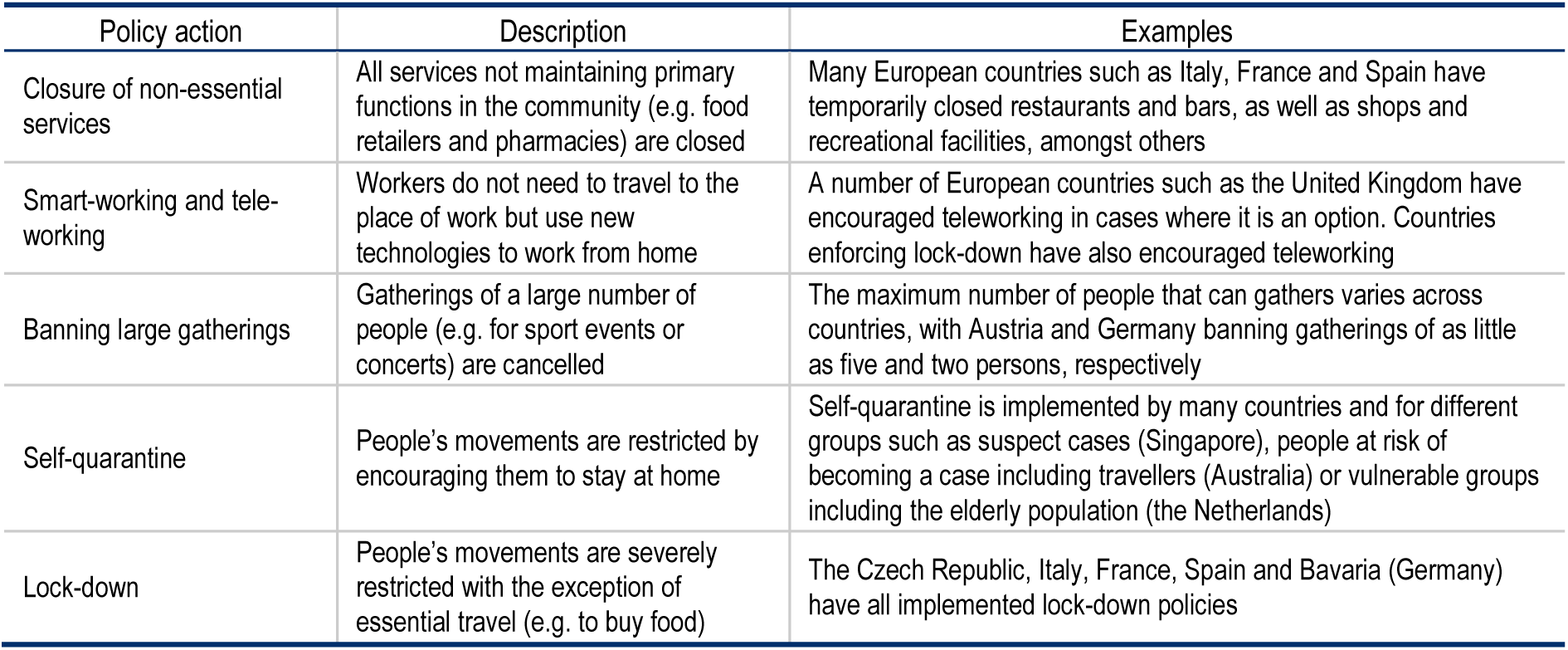
Examples of policies implemented by OECD countries to promote social distancing.

### School closures

School closures aim to decrease the number of contacts by school children, and thus reduce transmission of the disease throughout the community. There are two types of school closures: proactive school closures before any infection is associated with a school; and reactive school closures in response to a student, parent or staff member falling ill. School closures, either proactive or reactive, reduce influenza transmission but with a wide range of effectiveness varying between pandemics, cultures and setting (e.g. rural or urban), and depending on the level of transmission in schools and on the timing of the decision. School closures can, likely at best, reduce the peak attack rate by approximately 40% (Ferguson et al., 2006_[14]_; Cauchemez et al., 2009_[15]_) and delay the peak of the epidemic by a week or two (Rashid et al., 2015_[5]_; Bin Nafisah et al., 2018_[16]_). Reactive school closures may reduce influenza transmission by 7-15%, rarely up to 90-100% when transmission between children is assumed to be very influential (Rashid et al., 2015_[5]_). A study of novel H1N1 in New York found that reactive school closure reduce school-based transmission of influenza-like illness at school by 7% (Egger et al., 2012_[17]_). A modelling study estimated that school closures during an epidemic with a reproduction number of 2.5 would reduce the final attack rate from 65% to 60% (Milne et al., 2008_[18]_).

A number of factors modify the effectiveness of school closures. School closures are most effective for infections with limited rate of spread, when they are implemented in the early phases of an outbreak and when attack rates are higher in children than in adults (Jackson et al., 2014_[19]_). In addition, several studies suggest that measures to reduce out-of-school contacts should be implemented alongside school closures. Finally, school closures may need to be maintained throughout of the epidemic (Rashid et al., 2015_[5]_).

School closures have significant economic and social effects. Evidence shows that 16-45% of parents would need to take leave to supervise children at home; 16-18% of parents would lose income, and about 20% of households would have difficulty arranging childcare (Rashid et al., 2015_[5]_). School closures have high economic cost due to the loss of productivity of parents’ absenteeism from work. Based on data from the United Kingdom, economic modelling shows that 0.2-1% of GDP loss occurs due to school closure lasting the duration of a pandemic wave (Rashid et al., 2015_[5]_).

### Travel restrictions

To prevent or delay the entry of a disease into a country, it is common for policy-makers to implement several travel restrictions. Screening, for example, is common in airports during a disease outbreak, and may include thermal scans to identify passengers with a high external body temperature as well as health questionnaires to detect symptoms and possible exposure (by providing information on travel history) (Selvey, Antão and Hall, 2015_[20]_). Other restrictions include bans on (or advice to restrict) non-essential travel, voluntary or legally mandated isolation upon arrival into a new country as well as border closures (Mateus et al., 2014_[21]_; Australian Government, 2020_[22]_; Foreign & Commonwealth Office, 2020_[23]_)

Evidence to assess the effectiveness of travel restrictions are limited given they are frequently implemented alongside other countermeasures thereby making it difficult to ascertain causal effects (Mateus et al., 2014_[21]_). Consequently, the impact of national and international travel restrictions are typically measured using mathematical models. A systematic review by Mateus et al. (2014_[21]_) generalised findings from the literature related to influenza and concluded travel restrictions delay but do not prevent pandemics (e.g. delays up to 3-4 weeks when 90% of air travel is restricted in affected countries, or by two months if more restrictive measures are introduced). This finding aligns with experiences from other epidemic diseases. For example, a 60% reduction in airline passenger traffic from the Ebola affected region of West Africa was estimated to have delayed the spread of the disease to other continents by between 2-30 days (Poletto et al., 2014_[24]_). Regarding internal travel, a 2006 study found travel restrictions between cities in the United States during the 2001-02 influenza season delayed peak mortality by 16 days (Brownstein, Wolfe and Mandl, 2006_[25]_). The impact of internal travel restrictions have also been estimated for the COVID-19 outbreak, for example, Kucharski et al. (2020_[26]_) estimated that the introduction of travel control measures in Wuhan, China, reduced the median daily reproduction number from 2.35 to 1.05.

A major impact of travel restrictions is the flow-on effect this has on trade and business. As a result, travel restrictions dampen economic activity by reducing demand in tourism-dependent industries such as hotels, restaurants and aviation (Rashid et al., 2015_[5]_). In addition, screening may further stretch limited resources by isolating or quarantining travellers with symptoms unrelated to the disease of interest (Priest et al., 2015_[27]_).

### Contact tracing and quarantine

Contact tracing aims to identify, list and closely watch people who have been in contact with an infected person, even if they do not display symptoms. This helps the traced persons to get care and treatment early, and prevents further transmission of the virus (World Health Organization, 2017_[28]_). Quarantine (i.e. isolation) can be spent either at home, in hospital or specifically equipped structures.

There is limited data on the effectiveness of contact tracing and quarantine. Evidence that is available is largely based on modelling studies, which suggest quarantine decreases the peak case load, the attack rate and would also delay the peak. Household quarantine is potentially the most effective measure to reduce attack rates in the community, but only if compliance is high (Rashid et al., 2015_[5]_). Voluntary or self-isolation of infected people is moderately effective. For instance, voluntary quarantine of households with an infected individual may delay the peak of influenza by two to 26 days, and reduce the peak daily attack rate from 1.9% to 1.5%, or even down to 0.1%, depending on the associated interventions (e.g. such as treating infected people and applying prophylaxis to their households the day after the symptoms start, school closure) (Ferguson et al., 2006_[14]_). Modelling work simulating the isolation of cases and contacts in the case of the COVID-19 pandemic concluded that about 70% of cases had to be traced to successfully contain the outbreak, assuming a reproduction number of 2.5 (Hellewell et al., 2020_[29]_).

The effectiveness of this policy depends on numerous factors. First, it depends on whether infected people and their family members actually reduce their contact while they are ill. Another important factor is the time at which an infected individual becomes infectious. Isolation and quarantine is most effective in controlling the disease if the virus shedding starts after the onset of the clinical symptoms. For example, this was the case for SARS (the Severe Acute Respiratory Syndrome), a disease caused by another coronavirus as COVID-19, while, in the case of influenza, sick people are infectious during the incubation period (Fraser et al., 2004_[30]_).

Implementation of contact tracing and quarantine measures bears economic costs, and is associated with psychological, legal, and ethical issues. First, contact tracing requires substantial resources to sustain after the early phase of the epidemic since the number of infected people and contacts grow exponentially. While there is no obvious rationale for a routine use of contact tracing in the general population, it may be adapted in some circumstances (e.g. if there was an infected person on an aircraft) (Fong et al., 2020_[31]_). In addition, isolation is likely to cause distress and mental health problems, requiring additional services such as creating support lines and advice, helping people create plans, encouraging messages and calls and maintaining some routine (Lunn et al., 2020_[32]_).

### Public information campaigns

During an epidemic, policymakers can use campaigns to communicate with the public. These campaigns need to inform the public about the development of the epidemic and the risk it poses, with the aim of encouraging them to take the appropriate protective measures, such as hand washing or social distancing. In addition to saving lives, clear and timely information can also help preserve a country’s social, economic and political stability in the face of emergencies (World Health Organization, 2018_[33]_).

Especially in the case of COVID-19, which is a rapidly evolving situation with little knowledge about the disease, effective communication is crucial. Without it, the many unknowns can give space for rumours to develop and panic to set in (World Health Organization, 2018_[33]_). On the other hand, it is important to strike a balance between preventing panic and encouraging action. A study of the public perception of the influenza A/H1N1 (swine flu) outbreak in the United Kingdom found that few people changed their behaviour, and linked this to a believe that the outbreak had been exaggerated (Rubin et al., 2009_[34]_). The authors suggest that convincing the public that the threat is real may be a more pressing task for public health agencies than providing reassurance.

Evidence on the effectiveness of public information campaigns is limited. As mentioned above, the perceived risk can affect whether people change their hygiene behaviour, which means that the effectiveness of campaigns differs from one disease or epidemic to the next. Other studies have shown that the impact of campaigns can be increased by using trusted spokespeople like public health officials and through a role model effect from officials (Quinn et al., 2013_[35]_). The 2003 SARS outbreak hit Singapore in late February, prompting the government into action. An important element of Singapore’s containment strategy – in addition to the closure of schools and case isolation – was an integrated mass media campaign. This campaign included advertisements in the four languages (English, Chinese, Malay and Tamil) in newspapers and on television, a dedicated website, a toll-free hotline, booklets, posters and stickers. At the local level, town councils and other organizations organised discussion sessions and demonstrations for residents. A survey looking at whether the public followed the recommended behaviour found that adherence to the compulsory temperature checks was high: 85% of the respondents said that they monitored their temperature daily (Karan et al., 2007_[36]_).

### Environmental and personal hygiene

The evidence to date suggests COVID-19 is primarily spread from person-to-person via small respiratory droplets (GAVI Alliance, 2020_[37]_). The disease may also be transmitted through fomites, that is, objects that can carry infections (e.g. furniture) (Center for Disease Control and Prevention, 2020_[38]_).

In order to reduce the risk of transmission through fomites, policy-makers may require public and private spaces where infected individuals are likely, or known, to have frequented be cleaned and/or disinfected (e.g. schools, offices, day care centres) (Otter and Galletly, 2018_[39]_). This process is referred to as environmental decontamination (ED) and results in contamination levels that do not harm the health of individuals (Otter and Galletly, 2018_[39]_).

Given that the virus causing COVID-19 can remain on surfaces for an extended period of time, and that cleaning, as well as disinfectant, can reduce contamination levels, theoretically, ED can reduce transmission rates. Using these assumptions, a modelling study to estimate the impact of regular cleaning of high-touch surfaces in an office found the measure reduces the infection risk of influenza by 2.14% (Zhang and Li, 2018_[40]_).

The academic literature on environmental hygiene is limited with a recent systematic review identifying three studies (Xiao et al., 2020_[41]_). These studies all focus on younger children (i.e. those of school age) and do not accurately reflect ED efforts implemented during a pandemic. For example, one study measured the impact of disinfecting toys every two weeks (Ibfelt et al., 2015_[42]_), while another estimated the impact of bleach use in the home on respiratory illness (Casas et al., 2015_[43]_). For this reason, these studies should not necessarily be taken as evidence on the impact ED has on coronaviruses given SARS-CoV-2 can stay on a surface for up to three days.

The spread of COVID-19 has led countries to implement various ED policies. This is evidenced by the number of governments with online resources dedicated to best practice environmental and disinfection practices (e.g. (Australian Government Department of Health, 2020_[44]_; Center for Disease Control and Prevention, 2020_[38]_)).

In addition to environmental hygiene, individuals are encouraged to enhance their personal hygiene through hand washing, sneezing or coughing practices, and the use of protective facemasks. Various studies have shown these interventions can protect individuals by reducing their risk of getting infected (Figure 1) (Jefferson et al., 2011_[45]_). A study of the SARS outbreak in Hong Kong, China found that people who got infected were less likely to frequently have worn a face mask in public (odds ratio 0.36) or to have washed their hands 11 or more times per day (Lau et al., 2004_[46]_). A randomised trial during the influenza A(H1N1) pandemic showed a 35% to 51% reduction in the incidence of influenza-like illness when using both masks and exercising proper hand hygiene practices and cough etiquette (Aiello et al., 2010_[47]_). Similarly, a meta-analysis found that combining masks and hand hygiene reduced the risk of influenza infection by 27% (Wong, Cowling and Aiello, 2020_[48]_).

**Figure 1.**
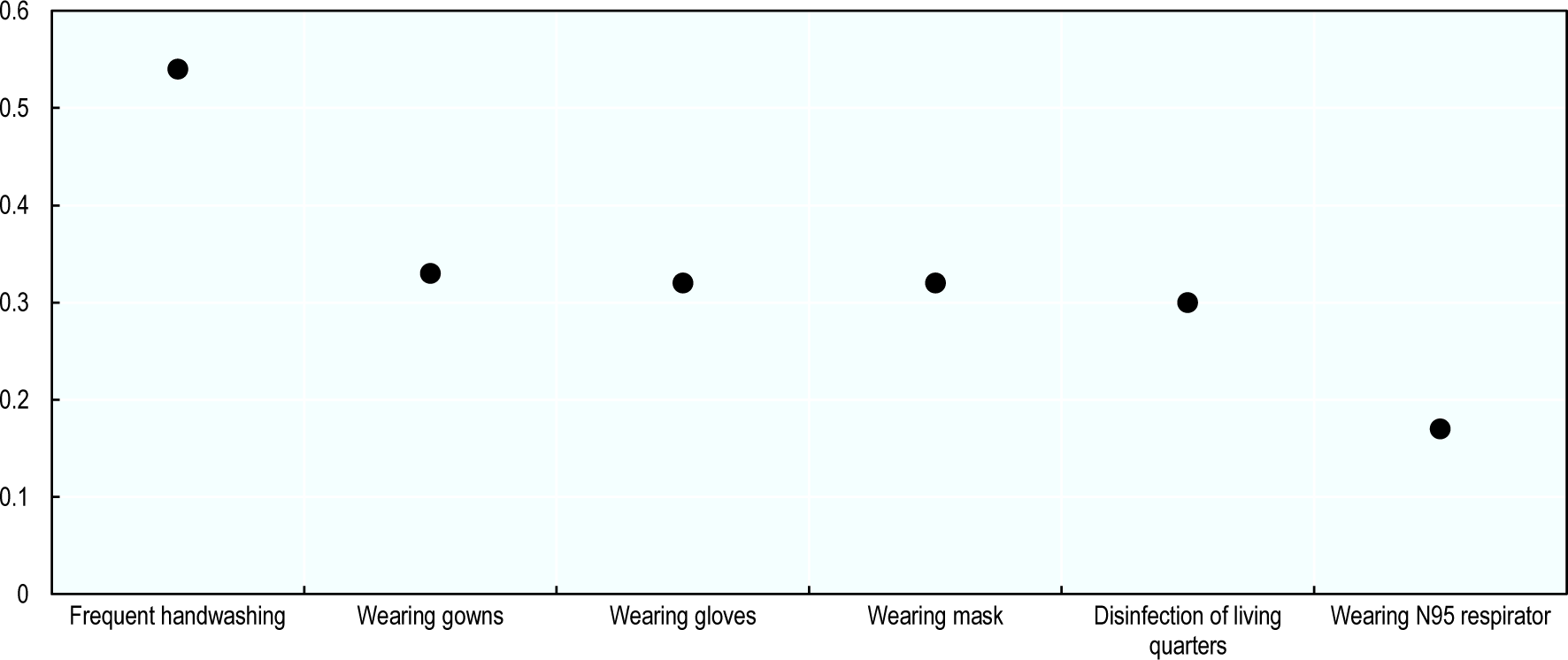
Impact of personal hygiene measures on respiratory viruses. Odds ratios Note: Results are reported as odds ratios (OR). An OR less (more) than one indicators a lower (higher) probability of contracting the virus. The lower the OR, the more effective the policy measure. Source: Adapted from Jefferson et al. (2011_[45]_)

**Figure 2.**
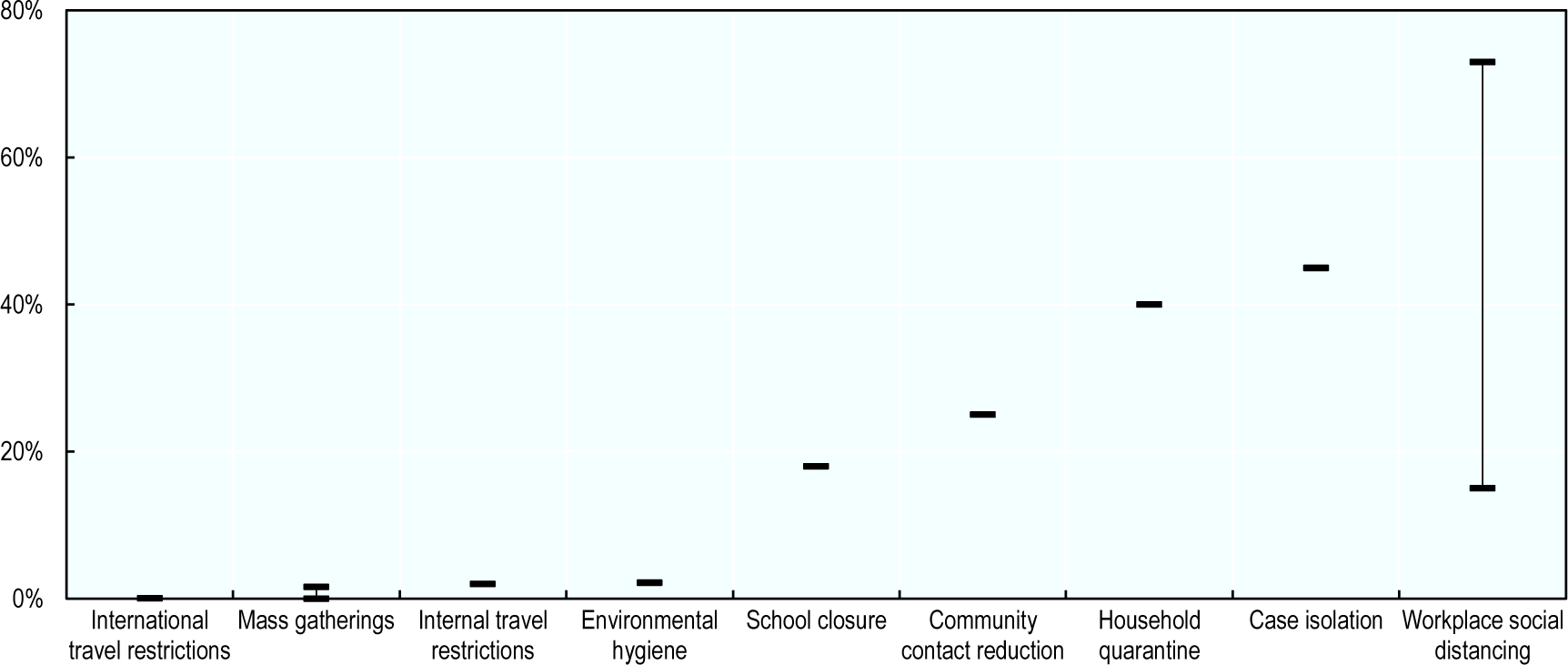
Impact of policy measures on influenza attack rates. Reduction in disease attack rates (%) Note: Not all policy measures are listed due to data availability. Source: OECD analyses on (Rashid et al., 2015_[5]_; Ahmed, Zviedrite and Uzicanin, 2018_[4]_; Mateus et al., 2014_[21]_; Milne et al., 2008_[18]_; Ferguson et al., 2006_[14]_).

**Figure 3.**
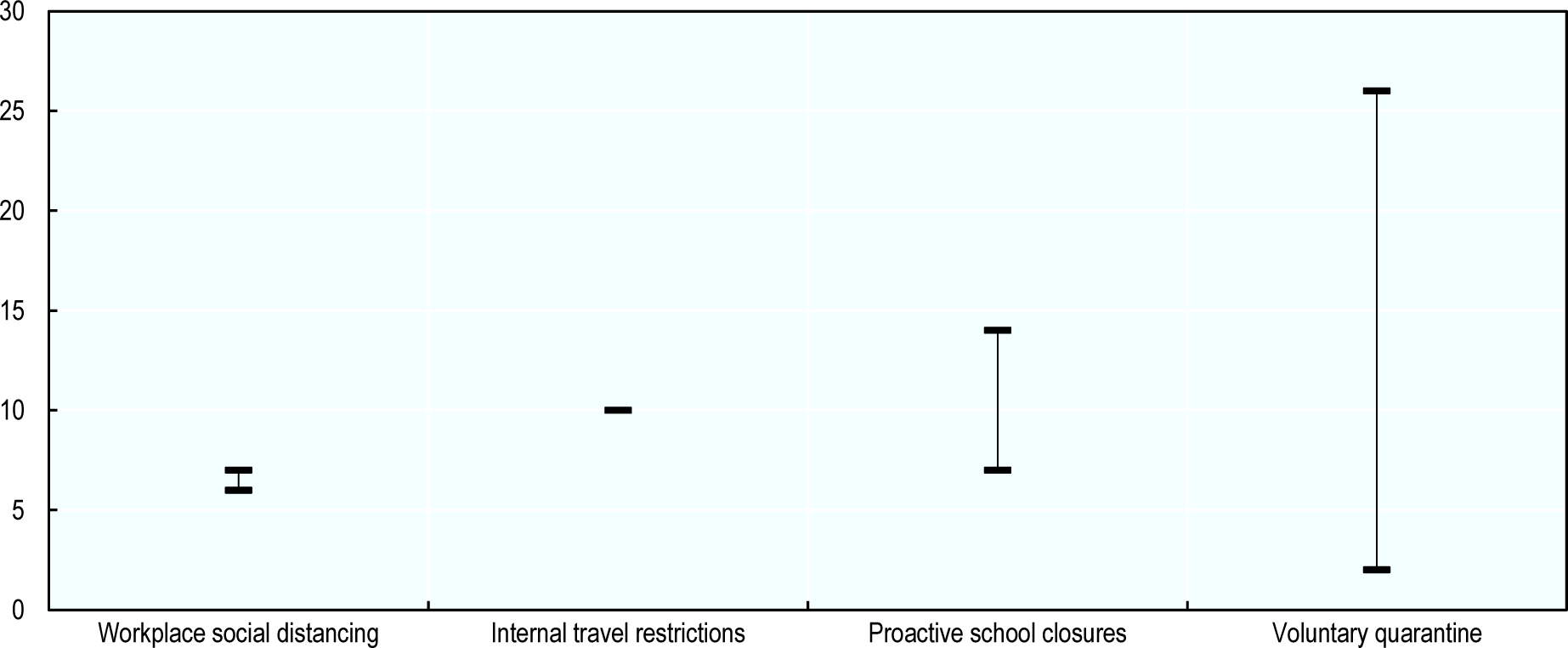
Impact of policy measures on the timing of influenza peak. Delay in the peak of the disease (in days) Note: Not all policy measures are listed due to data availability. Source: OECD analyses on (Rashid et al., 2015_[5]_; Ahmed, Zviedrite and Uzicanin, 2018_[4]_; Mateus et al., 2014_[21]_; Ferguson et al., 2006_[14]_).

### Summary of policy measures on key outcome measures

The impact of pandemic policy measures on outcome variables of interest are summarised in the figures below. Specifically, the impact on: 1) the attack rate, which represents the proportion of the population who are infected; and 2) the number of days the policy measure delays the peak of a disease.

Results from the analysis indicate that social distancing is the most effective measure for both reducing the attack rate as well as delaying the disease peak. For example, work place social distancing measures (such as working from home and workplace closures) can reduce the disease attack rate by between 23-73% (Ahmed, Zviedrite and Uzicanin, 2018_[4]_; Rashid et al., 2015_[5]_). Less effective measures include travel restrictions, in particular international restrictions, which is estimated to reduce the attack rate by just 0.02% (Mateus et al., 2014_[21]_).

Modelling studies consistently conclude that policy packages, as opposed to individual policies, are the most effective approach to reduce the impact of an epidemic (Lee, Lye and Wilder-Smith, 2009_[49]_). For example, Ferguson et al. (2006_[14]_) estimated that home quarantine reduces the overall attack rate by approximately 10% (i.e. from 27% to 24%), with this figure increasing to approximately 70% when adding school and workplace closures, effective border controls, and antiviral treatment and prophylaxis. A study from the United States found the reduction in the overall attack rate was 30 percentage points higher for a policy package including antiviral treatment and prophylaxis, quarantine, isolation and school closures (90% reduction in the attack rate) when compared to community and workplace social distancing only (60% reduction in the attack rate) (Halloran et al., 2008_[50]_). Finally, a modelling study from Milne et al. (2008_[18]_) estimated that a combination of school closures, case isolation, workplace non-attendance and limited community contact can reduce the final attack rate by 96% (i.e. from 55% to 2%) compared to between 12-45% (i.e. from 55% to 30-48%) when policies are implemented independently.

## Conclusions

This study outlines key NPIs used by various countries to combat the COVID-19 pandemic, including an assessment of their effectiveness. The objective of this analysis is two-fold: 1) to assist countries adapt their strategy to fight COVID-19, if necessary; and b) to inform transition strategies by identifying which NPIs to relax once the number of new cases falls and the impact of the disease lessens.

Based on an analysis of modelling studies, policy-makers are encouraged to implement policy packages to combat disease outbreaks such as COVID-19. Depending on the methodology and the policy package evaluated, these studies found comprehensive packages can reduce disease attack rates by at least 40%.

At the individual policy level, NPIs that reduce social interaction are most effective, in particular measures encouraging or requiring people to work at home, school closures and quarantine periods for those infected or potentially infected with the disease. These interventions, however, are associated with significant consequences including severely dampened economic activity and an increase in mental health issues. Less evidence is available to support interventions such as internal and international travel restrictions, bans on mass gatherings, environmental hygiene and public health campaigns.

Findings from this study broadly align with outcomes from Imperial College London’s modelling study, which found more stringent policies, such as school closures, have a greater impact on reducing disease transmission (Flaxman et al., 2020_[61]_).

Several limitations are associated with this study. First, this study relied on findings from a rapid review as opposed to a comprehensive systematic review of the literature. Second, only a selection of NPIs were analysed based on current discussions regarding potential policy actions to combat COVID-19. Third, findings from the analysis may not be directly applicable to combatting COVID-19 given the effectiveness of NPIs differ depending on the context in which they are implemented (e.g. population compliance) as well as the characteristics of the disease (e.g. reproduction number, incubation period, infection fatality rate). Finally, evidence to assess the impact of policy packages was reliant on modelling studies, which were often specific to a geographic region (primarily the United States and the United Kingdom).

## Data Availability

All the data used to feed the analysis is publicly available and sources have been referenced in the manyscript

## Acknowledgements

The views expressed in this paper are those of the authors and do not necessarily reflect those of the Organisation for Economic Co-operation and Development (OECD) or its member countries. The OECD program of work on public health is supported by a number of voluntary contributions by Ministries of Health or other national governmental institutions of OECD member countries or other key partners. No financial disclosures were reported by the authors of this paper.

## References

[14] Ahmed, F., N. Zviedrite and A. Uzicanin (2018), Effectiveness of workplace social distancing measures in reducing influenza transmission: A systematic review, BioMed Central Ltd., http://dx.doi.org/10.1186/s12889-018-5446-1.

[57] Aiello, A. et al.. (2010), “Mask Use, Hand Hygiene, and Seasonal Influenza-Like Illness among Young Adults: A Randomized Intervention Trial”, The Journal of Infectious Diseases, Vol. 201/4, pp. 491-498, http://dx.doi.org/10.1086/650396.

[12] Anderson, R. et al.. (2020), “How will country-based mitigation measures influence the course of the COVID-19 epidemic?”, The Lancet, Vol. 2019/20, pp. 1–4, http://dx.doi.org/10.1016/S0140-6736(20)30567-5.

[32] Australian Government (2020), COVID-19 (Coronavirus) and the Australian border, https://www.homeaffairs.gov.au/news-media/current-alerts/novel-coronavirus (accessed on 17 March 2020).

[54] Australian Government Department of Health (2020), Environmental cleaning and disinfection principles for COVID-19, https://www.health.gov.au/sites/default/files/documents/2020/03/environmental-cleaning-and-disinfection-principles-for-covid-19.pdf (accessed on 17 March 2020).

[5] Baum, S. (2020), COVID-19 Incubation Period: An Update, The New England Journal of Medicine (NEJM) Journal Watch, https://www.jwatch.org/na51083/2020/03/13/covid-19-incubation-period-update.

[7] Biggerstaff, M. et al.. (2014), “Estimates of the reproduction number for seasonal, pandemic, and zoonotic influenza: A systematic review of the literature”, BMC Infectious Diseases, Vol. 14/1, p. 480, http://dx.doi.org/10.1186/1471-2334-14-480.

[26] Bin Nafisah, S. et al.. (2018), “School closure during novel influenza: A systematic review”, Journal of Infection and Public Health, Vol. 11/5, pp. 657–661, http://dx.doi.org/10.1016/j.jiph.2018.01.003.

[22] Boddy, J., A. Young and P. O’Leary (2020), ‘Cabin fever’: Australia must prepare for the social and psychological impacts of a coronavirus lockdown, The Conversation, https://theconversation.com/cabin-fever-australia-must-prepare-for-the-social-and-psychological-impacts-of-a-coronavirus-lockdown-133353.

[23] Brooks, S. et al.. (2020), “The psychological impact of quarantine and how to reduce it: rapid review of the evidence”, The Lancet, Vol. 395/10227, pp. 912–920, http://dx.doi.org/10.1016/S0140-6736(20)30460-8.

[35] Brownstein, J., C. Wolfe and K. Mandl (2006), “Empirical Evidence for the Effect of Airline Travel on Inter-Regional Influenza Spread in the United States”, PLoS Medicine, Vol. 3/10, p. e401, http://dx.doi.org/10.1371/journal.pmed.0030401.

[16] Caley, P., D. Philp and K. McCracken (2008), “Quantifying social distancing arising from pandemic influenza”, Journal of the Royal Society Interface, Vol. 5/23, pp. 631–639, http://dx.doi.org/10.1098/rsif.2007.1197.

[53] Casas, L. et al.. (2015), “Domestic use of bleach and infections in children: a multicentre cross-sectional study”, Occupational and Environmental Medicine, Vol. 72/8, pp. 602–604, http://dx.doi.org/10.1136/oemed-2014-102701.

[25] Cauchemez, S. et al.. (2009), Closure of schools during an influenza pandemic, Elsevier, http://dx.doi.org/10.1016/S1473-3099(09)70176-8.

[48] Center for Disease Control and Prevention (2020), Environmental Cleaning and Disinfection Recommendations, https://www.cdc.gov/coronavirus/2019-ncov/community/organizations/cleaning-disinfection.html (accessed on 17 March 2020).

[1] Chen, N. et al.. (2020), “Epidemiological and clinical characteristics of 99 cases of 2019 novel coronavirus pneumonia in Wuhan, China: a descriptive study”, The Lancet, Vol. 395/10223, pp. 507–513, http://dx.doi.org/10.1016/S0140-6736(20)30211-7.

[13] Chen, S. et al.. (2020), “COVID-19 control in China during mass population movements at New Year.”, Lancet (London, England), Vol. 395/10226, pp. 764–766, http://dx.doi.org/10.1016/S0140-6736(20)30421-9.

[27] Egger, J. et al.. (2012), “The Effect of School Dismissal on Rates of Influenza-Like Illness in New York City Schools During the Spring 2009 Novel H1N1 Outbreak”, Journal of School Health, Vol. 82/3, pp. 123–130, http://dx.doi.org/10.1111/j.1746-1561.2011.00675.x.

[68] European Commission (2020), Coronavirus response: Transport measures, https://ec.europa.eu/transport/coronavirus-response_en.

[69] Federal Foreign Office (2020), Coronavirus and entry restrictions: 4 things travelers to Germany need to know, https://www.auswaertiges-amt.de/en/einreiseundaufenthalt/coronavirus (accessed on 20 March 2020).

[24] Ferguson, N. et al.. (2006), “Strategies for mitigating an influenza pandemic”, Nature, Vol. 442/7101, pp. 448–452, http://dx.doi.org/10.1038/nature04795.

[19] Ferguson, N. et al.. (2020), Impact of non-pharmaceutical interventions (NPIs) to reduce COVID-19 mortality and healthcare demand, Imperial College London, London, http://dx.doi.org/10.25561/77482.

[61] Flaxman, S. et al.. (2020), Estimating the number of infections and the impact of nonpharmaceutical interventions on COVID-19 in 11 European countries, Imperial College London, https://doi.org/10.25561/77731.

[41] Fong, M. et al.. (2020), “Nonpharmaceutical Measures for Pandemic Influenza in Nonhealthcare Settings—Social Distancing Measures”, Emerging Infectious Diseases, Vol. 26/5, http://dx.doi.org/10.3201/eid2605.190995.

[33] Foreign & Commonwealth Office (2020), Travel Advice against all non-essential travel: Foreign Secretary’s statement, 17 March 2020, https://www.gov.uk/government/news/travel-advice-foreign-secreatary-statement-17-march-2020 (accessed on 17 March 2020).

[40] Fraser, C. et al.. (2004), “Factors that make an infectious disease outbreak controllable”, Proceedings of the National Academy of Sciences of the United States of America, Vol. 101/16, pp. 6146–6151, http://dx.doi.org/10.1073/pnas.0307506101.

[47] GAVI Alliance (2020), What is COVID-19 and how is it spread?, https://www.gavi.org/vaccineswork/what-is-covid-19-and-how-does-it-spread (accessed on 17 March 2020).

[60] Halloran, M. et al.. (2008), “Modeling targeted layered containment of an influenza pandemic in the United States”, Proceedings of the National Academy of Sciences of the United States of America, Vol. 105/12, pp. 4639–4644, http://dx.doi.org/10.1073/pnas.0706849105.

[20] Hatchett, R., C. Mecher and M. Lipsitch (2007), “Public health interventions and epidemic intensity during the 1918 influenza pandemic”, Proceedings of the National Academy of Sciences of the United States of America, Vol. 104/18, pp. 7582–7587, http://dx.doi.org/10.1073/pnas.0610941104.

[39] Hellewell, J. et al.. (2020), “Feasibility of controlling COVID-19 outbreaks by isolation of cases and contacts”, The Lancet Global Health, Vol. 8, pp. e488–e496, http://dx.doi.org/10.1016/S2214-109X(20)30074-7.

[52] Ibfelt, T. et al.. (2015), “Effect of cleaning and disinfection of toys on infectious diseases and micro-organisms in daycare nurseries”, Journal of Hospital Infection, Vol. 89/2, pp. 109–115, http://dx.doi.org/10.1016/j.jhin.2014.10.007.

[17] Ishola, D. and N. Phin (2011), “Could influenza transmission be reduced by restricting mass gatherings? Towards an evidence-based policy framework”, Journal of Epidemiology and Global Health, Vol. 1/1, pp. 33–60, http://dx.doi.org/10.1016/j.jegh.2011.06.004.

[29] Jackson, C. et al.. (2014), The effects of school closures on influenza outbreaks and pandemics: Systematic review of simulation studies, Public Library of Science, http://dx.doi.org/10.1371/journal.pone.0097297.

[55] Jefferson, T. et al.. (2011), “Physical interventions to interrupt or reduce the spread of respiratory viruses”, The Cochrane database of systematic reviews 7, http://dx.doi.org/10.1002/14651858.CD006207.pub4.

[11] John Hopkins Medicine (2020), Coronavirus, Social Distancing and Self Quarantine, https://www.hopkinsmedicine.org/health/conditions-and-diseases/coronavirus/coronavirus-social-distancing-and-self-quarantine (accessed on 16 March 2020).

[46] Karan, K. et al.. (2007), “Emerging Victorious Against an Outbreak: Integrated Communication Management of SARS in Singapore Media Coverage and Impact of the SARS Campaign in Moving a Nation to be Socially Responsible”, Journal of Creative Communications, Vol. 2, pp. 383–403, http://dx.doi.org/10.1177/097325860700200307.

[36] Kucharski, A. et al.. (2020), “Early dynamics of transmission and control of COVID-19: a mathematical modelling study”, The Lancet Infectious Diseases, Vol. 0/0, http://dx.doi.org/10.1016/S1473-3099(20)30144-4.

[4] Lauer, S. et al.. (2020), “The Incubation Period of Coronavirus Disease 2019 (COVID-19) From Publicly Reported Confirmed Cases: Estimation and Application.”, Annals of internal medicine, http://dx.doi.org/10.7326/M20-0504.

[56] Lau, J. et al.. (2004), “SARS Transmission, Risk Factors, and Prevention in Hong Kong”, Emerging Infectious Diseases, Vol. 10/4, pp. 587–592, http://dx.doi.org/10.3201/eid1004.030628.

[71] Lee, V., C. Chiew and W. Khong (2020), “Interrupting transmission of COVID-19: lessons from containment efforts in Singapore”, http://dx.doi.org/10.1093/jtm/taaa039/5804843.

[59] Lee, V., D. Lye and A. Wilder-Smith (2009), “Combination strategies for pandemic influenza response - a systematic review of mathematical modeling studies”, BMC Medicine, Vol. 7/1, p. 76, http://dx.doi.org/10.1186/1741-7015-7-76.

[42] Lunn, P. et al.. (2020), “Using behavioural science to help fight the coronavirus”, ESRI working papers, No. 656, The Economic and Social Research Institute, Dublin, https://www.esri.ie/system/files/publications/WP656.pdf (accessed on 17 March 2020).

[18] Markel, H. et al.. (2007), “Nonpharmaceutical interventions implemented by US cities during the 1918-1919 influenza pandemic”, Journal of the American Medical Association, Vol. 298/6, pp. 644–654, http://dx.doi.org/10.1001/jama.298.6.644.

[31] Mateus, A. et al.. (2014), Effectiveness of travel restrictions in the rapid containment of human influenza: A systematic review, World Health Organization, http://dx.doi.org/10.2471/BLT.14.135590.

[28] Milne, G. et al.. (2008), “A Small Community Model for the Transmission of Infectious Diseases: Comparison of School Closure as an Intervention in Individual-Based Models of an Influenza Pandemic”, PLoS ONE, Vol. 3/12, p. e4005, http://dx.doi.org/10.1371/journal.pone.0004005.

[70] Ng, Y. et al.. (2020), “Evaluation of the Effectiveness of Surveillance and Containment Measures for the First 100 Patients with COVID-19 in Singapore — January 2–February 29, 2020”, MMWR. Morbidity and Mortality Weekly Report, Vol. 69/11, http://dx.doi.org/10.15585/mmwr.mm6911e1.

[21] OECD (2020), OECD Economic Outlook, Interim Report March 2020, OECD Publishing, Paris, https://dx.doi.org/10.1787/7969896b-en.

[65] OECD (2020), Supporting people and companies to deal with the Covid-19 virus: Options for an immediate employment and social-policy response, OECD, Paris, https://oecd.dam-broadcast.com/pm_7379_119_119686-962r78x4do.pdf.

[49] Otter, J. and T. Galletly (2018), “Environmental decontamination 1: what is it and why is it important?”, Nursing Times, https://cdn.ps.emap.com/wp-content/uploads/sites/3/2018/07/180627-Environmental-decontamination-1-what-is-it-and-why-is-it-important.pdf.

[34] Poletto, M. et al.. (2014), “Assessing the impact of travel restrictions on international spread of the 2014 west African Ebola epidemic”, Eurosurveillance, Vol. 19/42, http://dx.doi.org/10.2807/1560-7917.ES2014.19.42.20936.

[63] Premier of Victoria (2020), Statement From The Premier, https://www.premier.vic.gov.au/statement-from-the-premier-32/ (accessed on 22 March 2020).

[37] Priest, P. et al.. (2015), “Effectiveness of Border Screening for Detecting Influenza in Arriving Airline Travelers”, American Journal of Public Health, Vol. 105/S4, pp. S607–S613, http://dx.doi.org/10.2105/ajph.2012.300761r.

[66] Prime Minister of Australia (2020), Border restrictions, https://www.pm.gov.au/media/border-restrictions (accessed on 22 March 2020).

[45] Quinn, S. et al.. (2013), “Exploring Communication, Trust in Government, and Vaccination Intention Later in the 2009 H1N1 Pandemic: Results of a National Survey”, Biosecurity and Bioterrorism: Biodefense Strategy, Practice, and Science, Vol. 11/2, http://dx.doi.org/10.1089/bsp.2012.0048.

[15] Rashid, H. et al.. (2015), Evidence compendium and advice on social distancing and other related measures for response to an influenza pandemic, W.B. Saunders Ltd, http://dx.doi.org/10.1016/j.prrv.2014.01.003.

[44] Rubin, G. et al.. (2009), “Public perceptions, anxiety, and behaviour change in relation to the swine flu outbreak: cross sectional telephone survey”, BMJ, Vol. 339/b2651, https://doi.org/10.1136/bmj.b2651.

[30] Selvey, L., C. Antão and R. Hall (2015), “Entry screening for infectious diseases in humans”, Emerging Infectious Diseases, Vol. 21/2, pp. 197–201, http://dx.doi.org/10.3201/eid2102.131610.

[67] U.S. Department of State (2020), Current Outbreak of Coronavirus Disease 2019, https://travel.state.gov/content/travel/en/traveladvisories/ea/covid-19-information.html (accessed on 17 March 2020).

[62] UK Government (2020), Government announces further measures on social distancing, https://www.gov.uk/government/news/government-announces-further-measures-on-social-distancing (accessed on 22 March 2020).

[64] UNESCO (2020), School closures caused by Coronavirus (Covid-19), https://en.unesco.org/themes/education-emergencies/coronavirus-school-closures (accessed on 17 March 2020).

[10] van Doremalen, N. et al.. (2020), “Aerosol and Surface Stability of SARS-CoV-2 as Compared with SARS-CoV-1.”, The New England journal of medicine, p. NEJMc2004973, http://dx.doi.org/10.1056/NEJMc2004973.

[9] Wang, C. et al.. (2020), “Evolving Epidemiology and Impact of Non-pharmaceutical Interventions on the Outbreak of Coronavirus Disease 2019 in Wuhan, China”, medRxiv, p. 2020.03.03.20030593, http://dx.doi.org/10.1101/2020.03.03.20030593.

[58] Wong, V., B. Cowling and A. Aiello (2020), “Hand hygiene and risk of influenza virus infections in the community: a systematic review and meta-analysis”, http://dx.doi.org/10.1017/S095026881400003X.

[2] World Health Organization (2020), Coronavirus, https://www.who.int/health-topics/coronavirus (accessed on 16 March 2020).

[8] World Health Organization (2020), Coronavirus disease 2019 (COVID-19) Situation Report – 46, https://www.who.int/docs/default-source/coronaviruse/situation-reports/20200306-sitrep-46-covid-19.pdf?sfvrsn=96b04adf_2 (accessed on 16 March 2020).

[6] World Health Organization (2018), Influenza (Seasonal), https://www.who.int/news-room/fact-sheets/detail/influenza-(seasonal) (accessed on 16 March 2020).

[43] World Health Organization (2018), Managing epidemics: Key facts about major deadly diseases.

[38] World Health Organization (2017), WHO Contact tracing, https://www.who.int/features/qa/contact-tracing/en/ (accessed on 17 March 2020).

[51] Xiao, J. et al.. (2020), “Nonpharmaceutical Measures for Pandemic Influenza in Nonhealthcare Settings—Personal Protective and Environmental Measures”, Emerging Infectious Diseases, Vol. 26/5, http://dx.doi.org/10.3201/eid2605.190994.

[3] Xu, X. et al.. (2020), “Clinical findings in a group of patients infected with the 2019 novel coronavirus (SARS-Cov-2) outside of Wuhan, China: Retrospective case series”, The BMJ, Vol. 368, http://dx.doi.org/10.1136/bmj.m606.

[72] Zastrow, M. (2020), “South Korea is reporting intimate details of COVID-19 cases: has it helped?”, Nature, http://dx.doi.org/10.1038/d41586-020-00740-y.

[50] Zhang, N. and Y. Li (2018), “Transmission of influenza a in a student office based on realistic person-to-person contact and surface touch behaviour”, International Journal of Environmental Research and Public Health, Vol. 15/8, http://dx.doi.org/10.3390/ijerph15081699.

